# Viral genomes reveal patterns of the SARS-CoV-2 outbreak in Washington State

**DOI:** 10.1101/2020.09.30.20204230

**Authors:** Nicola F. Müller, Cassia Wagner, Chris D. Frazar, Pavitra Roychoudhury, Jover Lee, Louise H. Moncla, Benjamin Pelle, Matthew Richardson, Erica Ryke, Hong Xie, Lasata Shrestha, Amin Addetia, Victoria M. Rachleff, Nicole A. P. Lieberman, Meei-Li Huang, Romesh Gautom, Geoff Melly, Brian Hiatt, Philip Dykema, Amanda Adler, Elisabeth Brandstetter, Peter D. Han, Kairsten Fay, Misja Ilcisin, Kirsten Lacombe, Thomas R. Sibley, Melissa Truong, Caitlin R. Wolf, Michael Boeckh, Janet A. Englund, Michael Famulare, Barry R. Lutz, Mark J. Rieder, Matthew Thompson, Jeffrey S. Duchin, Lea M. Starita, Helen Y. Chu, Jay Shendure, Keith R. Jerome, Scott Lindquist, Alexander L. Greninger, Deborah A. Nickerson, Trevor Bedford

## Abstract

The rapid spread of SARS-CoV-2 has gravely impacted societies around the world. Outbreaks in different parts of the globe are shaped by repeated introductions of new lineages and subsequent local transmission of those lineages. Here, we sequenced 3940 SARS-CoV-2 viral genomes from Washington State to characterize how the spread of SARS-CoV-2 in Washington State (USA) was shaped by differences in timing of mitigation strategies across counties, as well as by repeated introductions of viral lineages into the state. Additionally, we show that the increase in frequency of a potentially more transmissible viral variant (614G) over time can potentially be explained by regional mobility differences and multiple introductions of 614G, but not the other variant (614D) into the state. At an individual level, we see evidence of higher viral loads in patients infected with the 614G variant. However, using clinical records data, we do not find any evidence that the 614G variant impacts clinical severity or patient outcomes. Overall, this suggests that at least to date, the behavior of individuals has been more important in shaping the course of the pandemic than changes in the virus.

**One Sentence Summary:** Local outbreak dynamics of SARS-CoV-2 in Washington State (USA) were driven by regionally different mitigation measures and repeated introductions of unique viral variants with different viral loads.

## Introduction

After its emergence near the end of November or beginning of December 2019 in Wuhan, China, SARS-CoV-2 rapidly spread around the world (*1*). In the United States, the first reported case of COVID-19, the disease caused by SARS-CoV-2, was found in Washington State on January 19, 2020 in a traveler who returned from China 4 days earlier. Until the end of February, no additional cases of COVID-19 were reported in Washington State.

At the end of February, however, a case of COVID-19 was reported in Snohomish County, the same county where the initial case was reported. This case had no known travel history and constitutes the first reported case of community transmission in Washington State (*2*). While genetically closely related to the initial case, the later sequenced cases share a common ancestor in early February and have been reported to likely be due to an independent introduction (*2*).

After these initial introductions, SARS-CoV-2 has been introduced repeatedly into Washington State from different parts of the globe. Viruses introduced later differ genetically from those introduced earlier, most notably in one amino acid in the spike protein, which facilitates viral entry and includes the receptor-binding domain. Since its first occurrence, this amino acid substitution from aspartate (D) to glycine (G) at position 614 of the Spike protein increased in relative frequency around the world (visible at: https://nextstrain.org/ncov/global?c=gt-S_614) and now represents the vast majority of all new cases of COVID-19 (*3–5*). This increase in relative frequency of the 614G variant has been proposed to be due to higher transmissibility of the 614G variant over the 614D variant (*4, 6*). A modest increase in viral load has been observed in patients infected with the 614G variant (*4, 7*). Recently, multiple *in vitro* studies in human cell lines found a 3-9 fold increase in infectivity of the 614G variant (*5, 8, 9*). However, it remains unclear whether the population level trends are due to higher transmissibility of the virus, or simply due to founder effects, i.e. owing to strong bottlenecks when SARS-CoV-2 spread globally, as the D614G variant got a start early on in the European COVID-19 epidemic and spread from Europe to the rest of the world.

Washington State differs regionally, from more densely populated areas at the coast, to more sparsely populated areas inland. We here focus on differences between the spread on lineages of 614D and 614G in the context of regional differences within Washington State. Extensive local spread of SARS-CoV-2 was first detected in King County, which includes the city of Seattle. King County was also the first region in the state to take action to curb the spread of SARS-CoV-2, including several large companies in the area mandating work from home in early March 2020 (*10*). After a statewide lockdown, new cases began to fall in the whole state, except for Yakima County, where cases peaked substantially later than in the rest of the State.

Using viral genetic sequence data isolated from patients in Washington State between February and July 2020, we test the impact of temporal differences in county level mobility trends, as well as the role of introductions from outside the state in driving case loads. We additionally investigate potential transmissibility differences by comparing viral loads using cycle thresholds for viral quantification. Lastly, we investigate whether the D614G amino acid substitution leads to more severe disease in patients infected with SARS-CoV-2.

### Outbreak in Washington State caused by repeated introductions and shaped by temporal differences in mobility reductions

We sequenced 3940 viruses from Washington State collected between February and July 2020 and use these sequences alongside other publicly available sequences from elsewhere in the world to characterize transmission dynamics. We observe SARS-CoV-2 entered Washington State from different parts of the world and subsequently spread locally, evident as clusters of genetic similar Washington State viruses in the global phylogeny (Fig. 1A). An early February introduction of a 614D variant (*2, 11*) fueled much of the early outbreak in March and April, but this lineage was supplanted through multiple introductions of 614G and past April the majority of viruses are 614G (Fig. 1).

**Fig 1.**
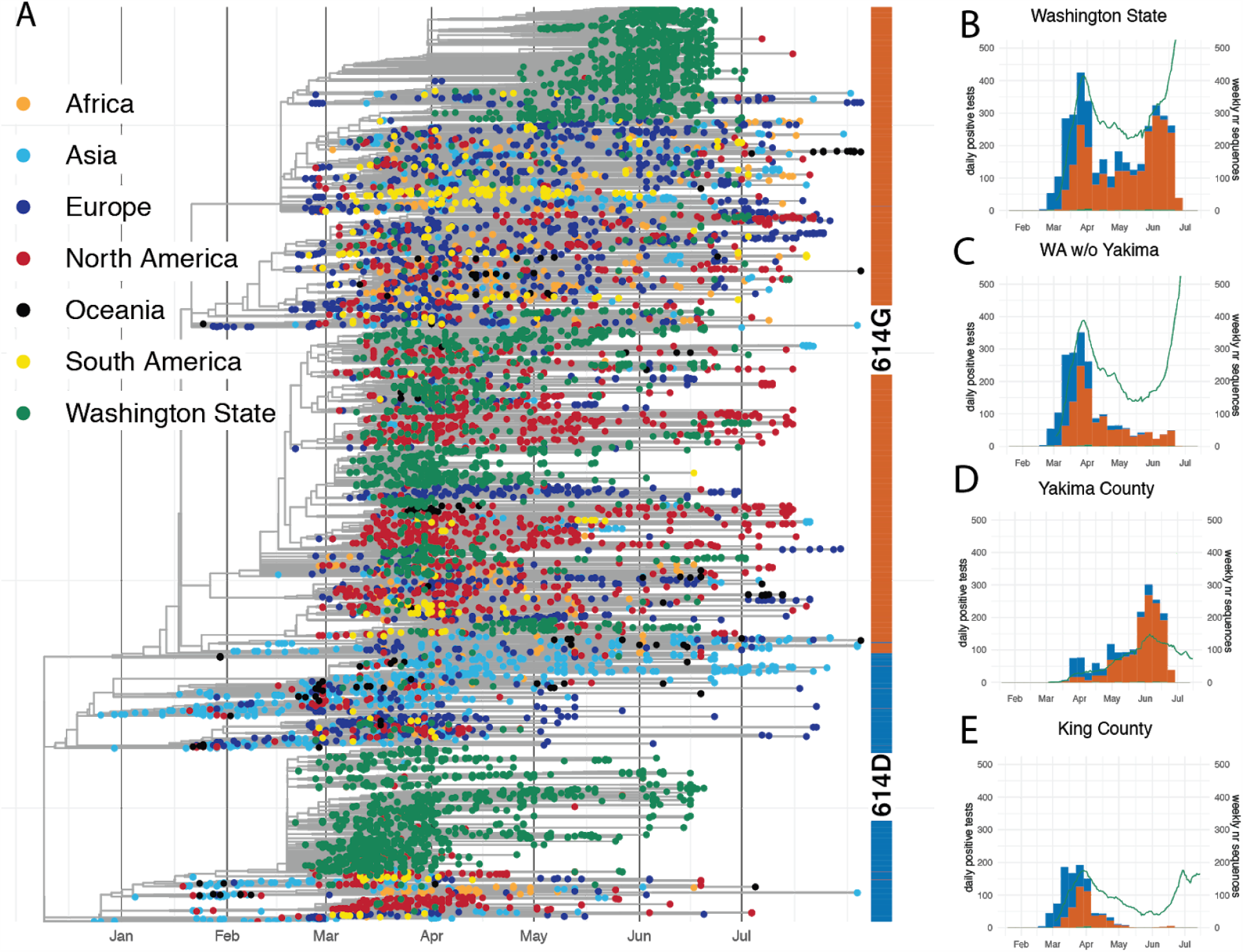
SARS-CoV-2 phylogeny highlighting D614G split and cases through time in Washington State. (A) Phylogenetic tree of 10,051 sequences from Washington State and around the world. Tips are colored based on sampling location. This is a time-calibrated phylogeny with time shown on the x-axis. The split between 614D sequences (blue) and 614G (orange) sequences is shown as a bar to the right of the phylogeny. (B-E) Confirmed cases and genetic makeup of SARS-CoV-2 across Washington State and individual counties. The green line shows a 7 day moving average of daily confirmed cases. The bar plots show weekly sequenced cases in our dataset. Cases due to the 614D variant are shown in blue and cases due to the 614G variant are shown in orange.

To analyse the introduction and local spread of SARS-CoV-2 in Washington State, we first split these sequences into different local transmission clusters, which we define as groups of sequences that originate from a single introduction into Washington State. To do so, we use a parsimony based clustering approach, considering Washington State and everything outside Washington State as the two possible locations for parsimony clustering. The local transmission clusters obtained are shown at https://nextstrain.org/groups/blab/ncov/wa-phylodynamics?c=cluster_size and their size distribution and D614G makeup shown in Figure S1. We then use these local transmission clusters to analyse the spread of SARS-CoV-2 in the state using two phylodynamic approaches. First, we estimate the effective reproduction number (*R*_*e*_) using a birth-death approach (*12*), where we treat each individual local transmission cluster as independent observation of the same underlying population process (*13*). Next, we estimate effective population sizes over time and the degree of introductions using a coalescent skyline approach (*14*). To do so, we assume that all sequences that cluster together are the result of local transmission and each individual cluster is the result of one introduction into Washington State. We then model the whole process as a structured coalescent process (*15, 16*) where we assume to know the migration history based on the previous clustering (see Methods and Material for details). In contrast to the birth-death model, the coalescent conditions on sampling, meaning that the information about population level trends comes from the phylogenetic tree itself and not from the number of sequences through time.

We perform these phylodynamic analyses for a random subsample of 1500 samples from all Washington counties except for Yakima County as well as for the 614D (500 sequences) and 614G (1000 sequences) lineages separately. Additionally, we performed the same analysis using 750 sequences from Yakima County only. After an initial introduction (*2*), the number of cases grew rapidly (Fig. 2A). As expected, growth in confirmed cases is mirrored in phylodynamic estimates of viral effective population size (Fig. 2A). Additionally, we observe maximal transmission intensity at the end of February when *R*_*e*_ is between 2 and 3 (Fig. 2B). This is consistent with other estimates of the effective reproduction number of SARS-CoV-2 during early phases of an epidemic when control measures are not in place (*17–19*).

**Fig 2.**
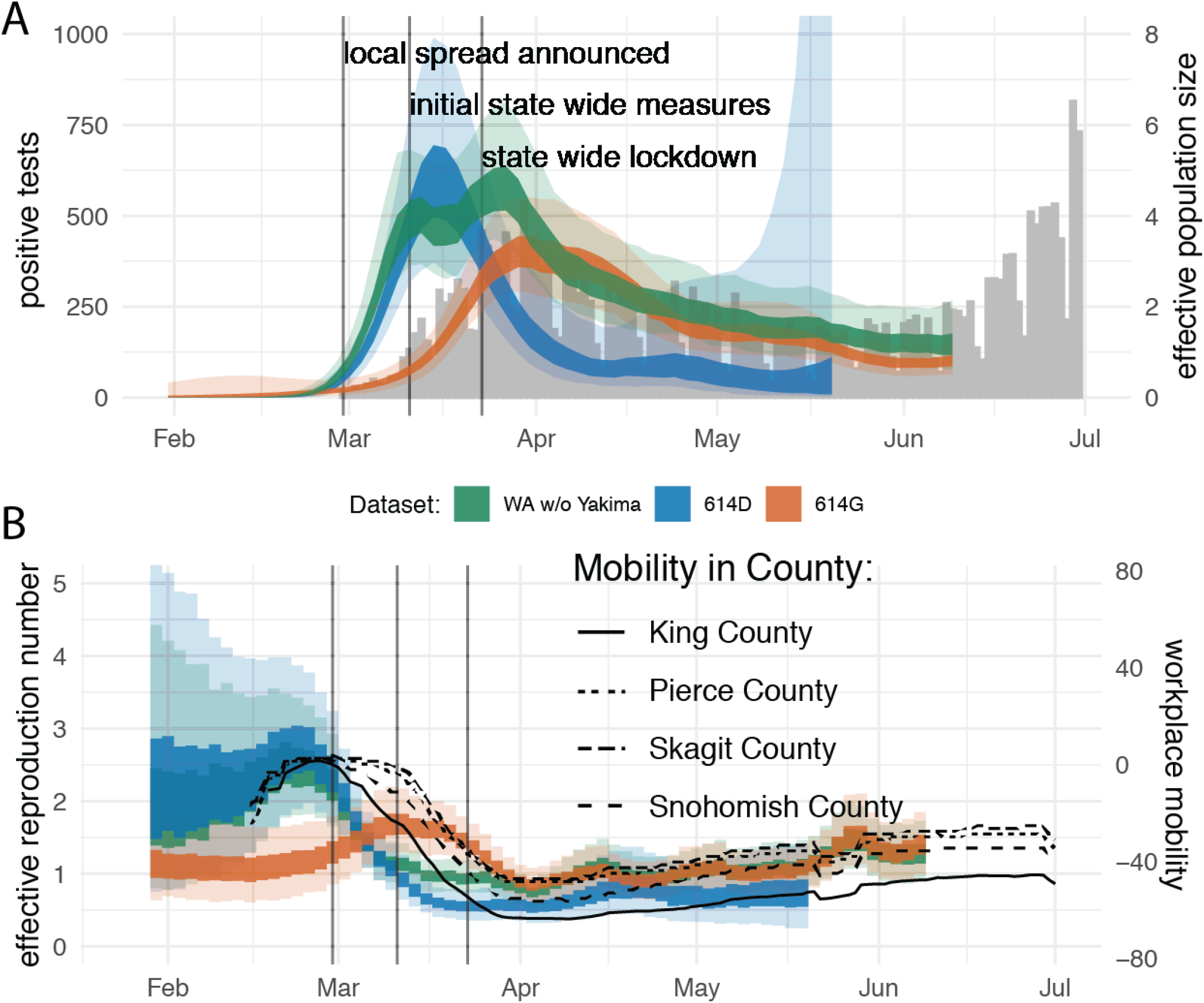
Regional dynamics of SARS-CoV-2 in Washington State inferred from confirmed cases and pathogen genomes. (A) Estimates of effective population sizes for the outbreak in Washington State (green interval), as well as for 614D (blue interval) and 614G (orange interval) individually compared to confirmed cases in the state (gray bars). The inner band denotes 50% highest posterior density (HPD) interval and the outer band denotes the 95% HPD interval. (B) *R*_*e*_ estimates using a birth death approach for the same groups as in (A). The *R*_*e*_ estimates are compared to Google workplace mobility data for King, Pierce, Skagit and Snohomish Counties shown as black solid and dashed lines. Workplace mobility is represented as a 7 day moving average.

Around the time when community spread in King County was announced on February 29, 2020, we observe decreased occupancy of workplaces according to Google mobility data (Fig. S2) (*20*). This reduction in workplace mobility occurred earlier in King County, compared to other regions of the state that had little or no reported cases at the time (Fig. S2). This is consistent with several businesses starting to institute measures, such as work-from-home policies, at the beginning of March (*10*). This reduction in mobility in King County coincided with a reduction in the effective reproduction number of 614D cases in the state (Fig. 2B). By the time initial statewide measures were taken on the 11th of March, cases of 614D had almost peaked and were starting to decline while overall cases were approximately constant or still increasing (Fig. 2A).

Cases of 614G were still increasing and peaked a little over a week later than cases of 614D (Fig. 1 and 2A). This was around the time when the statewide lockdown order came into effect on March 24, 2020. While cases of 614D were initially mostly located around Seattle, cases of 614G were more widespread throughout the state. Viruses sampled from cases in Pierce County and in the counties north of King County mostly harbored the 614G variant (Fig. 1C). Changes in the effective reproduction number of 614G coincided with changes in mobility outside of King County (Fig. 2B). An alternative phylodynamic method using a coalescent approach yields highly similar results (Fig. S3).

Yakima County was the other county in the state (besides King County) with a significant number of 614D cases later in the epidemic (Fig. 1D). The outbreak there happened later than the first large outbreak in King and neighboring counties. Additionally, the trend in cases in Yakima County became increasingly decoupled from workplace mobility as measured by cellphone movement for reasons likely associated with a large population of essential workers in the agricultural sector and seasonal worker migration poorly captured in mobility metrics (Fig. S4) (*21, 22*).

We next investigated the importance of introductions in driving the outbreak in Washington State. To do so, we estimated the relative contribution of introductions compared to local transmission following the coalescent approach introduced above. In short, we use the estimated changes in effective population sizes over time and the estimated rates of introduction to compute the percentage of new cases in the state due to introductions (see Material and Methods for details).

We estimate the percentage of new cases due to introductions in Washington State (excluding Yakima County) to be below 10% initially and to then have increased to about 10% by the middle of March through early April (Fig. 3). As a reference, the US instituted a travel ban for non-residents coming from China on February 2, 2020, and a travel ban from Europe effective March 16, 2020. Increases in the proportion of introductions of the overall cases can either be driven by a reduction in the local transmission rate and or an increase in the rate of introduction.

**Fig 3.**
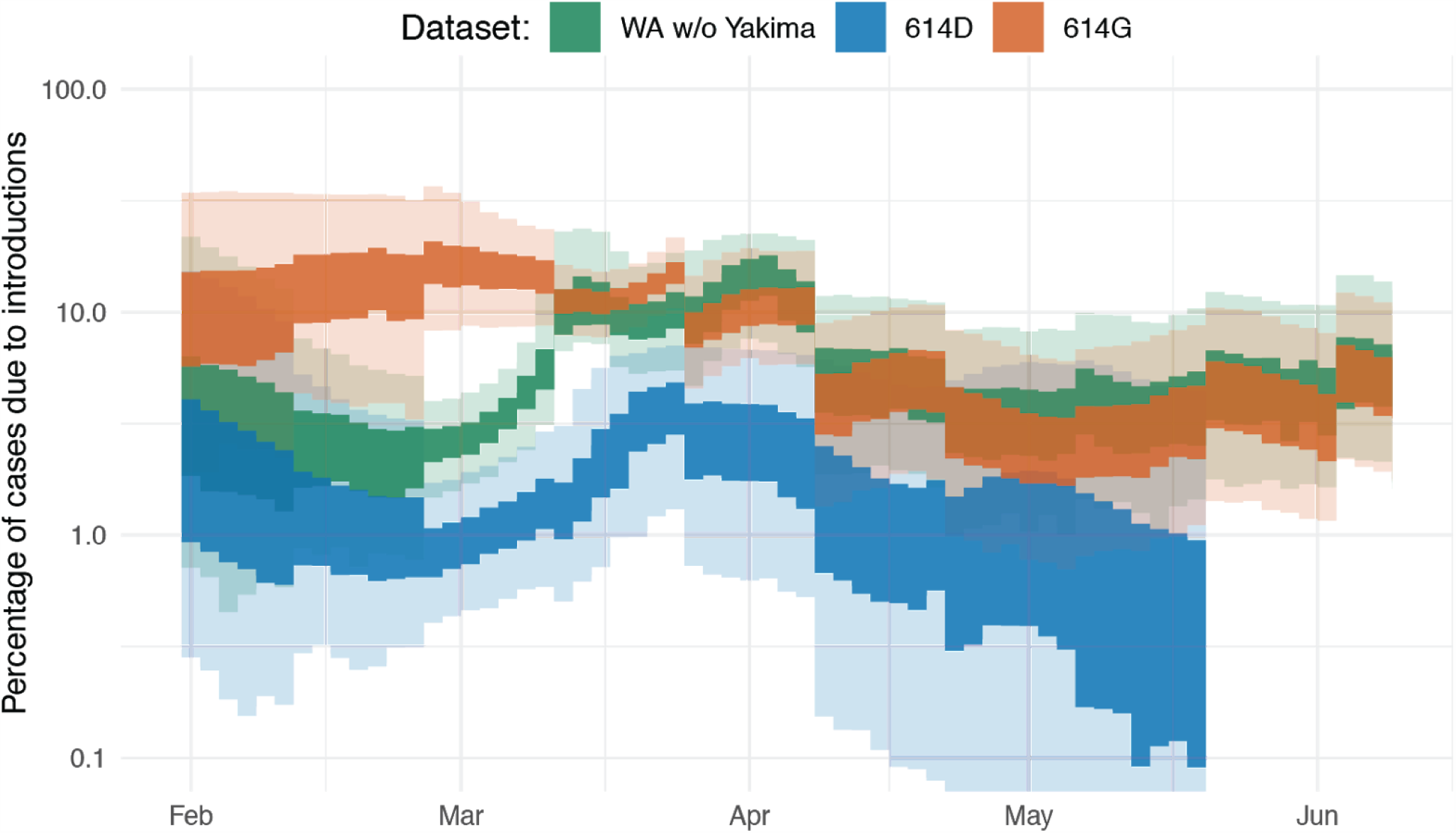
Phylogenetic estimate of the percentage of introductions of the overall cases. Proportions are estimated as the relative contribution of introductions to the overall number of infections using the multi-tree coalescent. Proportions are shown for the outbreak in Washington State (green interval), as well as for 614D (blue interval) and 614G (orange interval). The inner area denotes the 50% HPD interval, the outer area denotes the 95% HPD interval.

These introductions were unevenly distributed across the different clades 614D and 614G (Fig. 3) (*6, 23*). The proportion of introduced 614G cases is substantially greater than the proportion of introduced 614D cases. We estimate the percentage of introduced 614D cases to be below 3% during the whole outbreak. On the other hand, we infer the percentage of introduced 614G cases to have been over 10% until the beginning of April. This means that a substantially higher fraction of 614G cases were caused by introductions than in 614D cases. This is expected considering that cases of 614G were much more widespread outside of China (Fig. 1A), including in areas with relatively strong travel patterns to Washington State during the epidemic, such as New York State.

We next tested whether the percentage of new cases caused by introductions are reasonable given the number and size distribution of local transmission clusters. To do so, we simulated local transmission clusters where 0.1%, 1% or 10% of all infections are caused by novel introductions. We find that the observed patterns in transmission cluster size distributions fall between the simulated patterns for 1% and 10% of all infections having been caused by introductions (Fig. S5).

Overall, it appears that population level changes in Washington State in relative frequencies of the two lineages can be explained by differences in timing of measures to curb the spread of SARS-CoV-2 on a county level and by repeated introductions of 614G. Although a parsimonious explanation of observed dynamics, this does not preclude 614G having a higher transmission rate relative to 614D. Additionally, these population level trends are impacted by many confounding factors that are not directly related to the virus itself. We therefore next move to investigate whether we can observe differences between patients infected with viruses from either lineage on an individual level.

### D614G leads to higher viral load, without apparent effects on virulence

We tested for differences in viral loads between patients infected with either the 614D or the 614G viral variants by comparing cycle threshold (Ct) values. Ct values are inversely correlated with viral load, and differences in Ct values between these two variants have been reported previously (*4, 7*). To test this, we analyzed 1770 SARS-CoV-2 sequenced samples from Washington State for which we had access to Ct values. We only used genomes sampled between February and April 2020, when both lineages were circulating in Washington State.

Of these 1770 genomes, 1128 genomes were from patients referred by a healthcare provider for nasopharyngeal swab testing to the University of Washington (UW) Virology laboratory. 550 genomes were from samples collected by the Washington Department of Health (WA DOH), and 92 samples were from self-collected nasal swabs mailed in for testing as part of the Seattle Coronavirus Assessment Network (SCAN). During this time period, UW Virology used multiple platforms for PCR testing (Fig. S6). Since it is difficult to compare Ct across primer sets and platforms (*24*), we mainly focus on samples amplified with the most common primer set: N1, N2 (*n*=879), although analyses of results using ORF1ab primers (*n*=229) were also conducted.

We found that patients infected with viruses with the 614G substitution had lower Ct values (higher viral load) than those infected with 614D viruses in all three collection channels (Figs. 4A, S7). This difference was significant by Wilcoxon Rank Sum Test in samples from UW Virology (N1, N2 primers: median Δ = 1.5 cycles, *p* = 1.5e-12, ORF1ab primers: median Δ = 2.5 cycles, *p* = 0.0012) and WA DOH (median Δ = 1.5 cycles, *p* = 0.014), but not when only using the samples from SCAN, where we had far fewer samples (median Δ = 2.1 cycles, *p* = 0.077) (Figs. 4A, S7).

**Fig 4.**
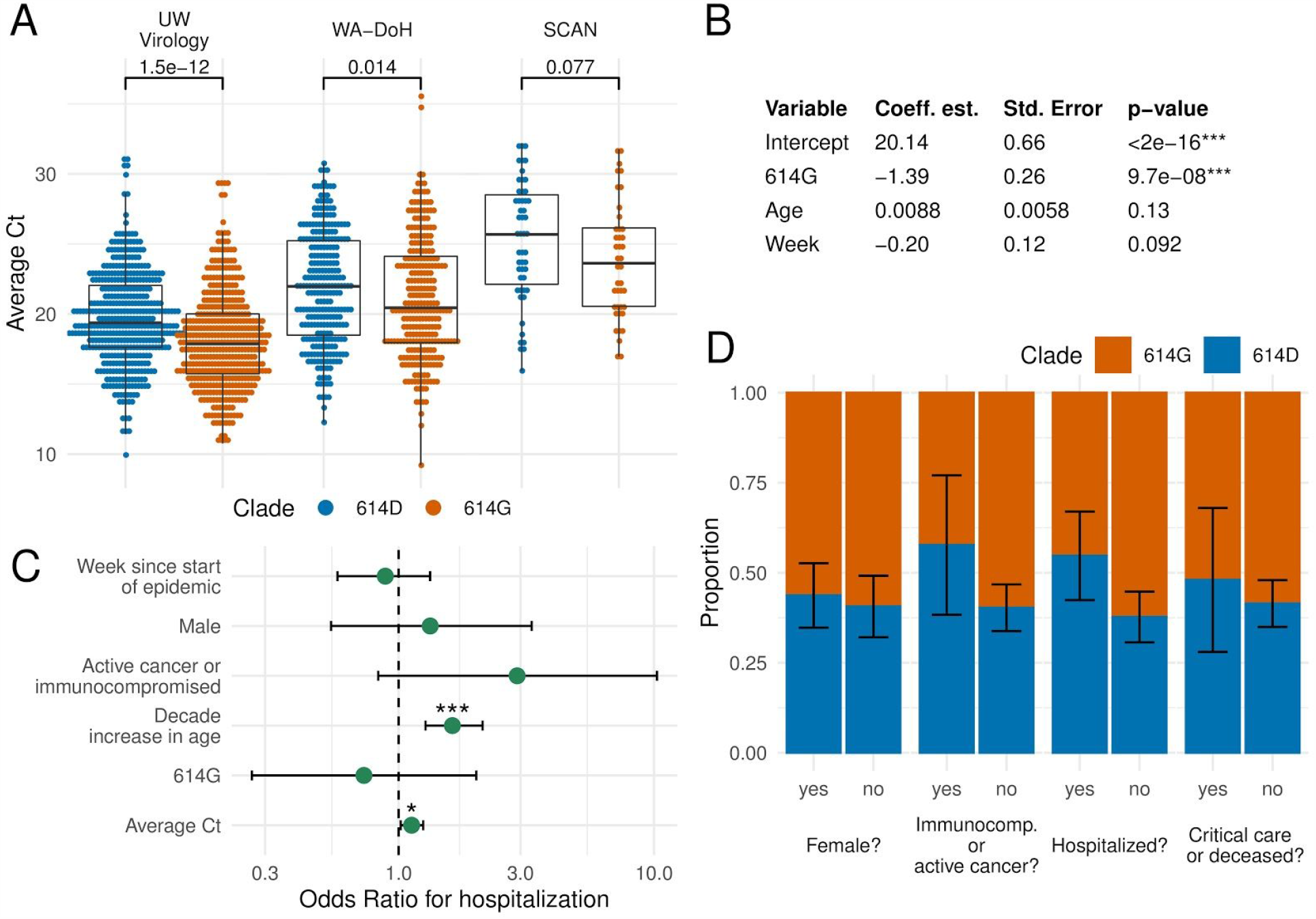
Factors affecting viral load and disease severity at an individual level. **A** Comparison between cycle threshold (Ct) values for viruses from the 614D and 614G clade. **B** GLM analysis of Ct values using variant, age, and sampling week as predictors. **C** Odds ratios of being hospitalized given being infected with SARS-CoV-2. Error bars show 95% CI, corrected for multiple hypothesis testing using a Bonferroni Correction. **D** Proportion of viruses in 614D and 614G clades grouped by sex, immunocompromised status, hospitalization, and severe outcome (requiring critical care or death). Proportion was calculated as the mean of a binary clade variable; error bars show standard error of the mean.

We next tested whether factors other than D614G variant predict Ct values. To do so, we applied a generalized linear model (GLM), assuming normally distributed Ct values, to the UW Virology samples using variant, patient age, and calendar week of sampling as potential predictors of Ct values. If later in the epidemic, carriers of SARS-CoV-2 are detected at an earlier stage of their infection, the measured Ct values would be impacted purely based on how long after infection a patient has been tested (*25–28*). We include sampling week as a potential predictor of Ct values to potentially correct for this.

We find that D614G variant is the best predictor of Ct values, with 614G having a Ct value that is, on average, 1.4 cycles lower than for 614D (N1, N2 primers) (Fig. 4B) when controlling for age and sampling week. This difference in Ct translates to a 0.42 log_10_ increase in viral load (95% CI: 0.26-0.57 log), assuming the standard curve is linear in this region. With ORF1ab primers, we observe similar coefficients and significance in the GLM (Fig. S7); however, the residuals are not normally distributed, suggesting the model fits poorly with ORF1ab primers.

We also found a difference in age of people infected between the two lineages (Fig. S8). In samples from UW Virology, the average age of patients infected with viruses from the 614D and 614G lineages were 56.6 and 52.4, respectively (*p* = 5.8e-04, Student’s *t*-test). In SCAN samples, the average age of patients was 45.8 for 614D and 38.4 for 614G (*p* = 0.088). Age differences may be caused by increased testing, resulting in detection of less severe, younger cases later in the epidemic when 614G was more prevalent (Figs. 1 and 2). However, we tested this hypothesis in a GLM with week of sample collection and D614G variant as potential predictors of age. Individuals with 614G variant were 3.5 years younger on average (*p* = 0.0098) while sample week was not a significant predictor of age (*p* = 0.20) (Fig. S8). A skew towards younger individuals is consistent with either a more transmissible virus or with more severe infection as this would result in a larger fraction of younger patients seeking testing. However, the absolute difference in age of infection is still small.

For 248 of the 1128 sequences from patients referred for SARS-CoV-2 testing by a healthcare provider, we had access to additional clinical information. 104 of these patients were infected with viruses from the 614D clade, and 144 patients were infected with viruses from the 614G clade. We used data from electronic health records to examine if differences in Ct values hold after correcting for additional potentially confounding factors. We performed the same GLM analysis as above, but included additional potential predictors, such as sex, active cancer or immunocompromised status, hospitalization, and whether a patient required intensive care or died.

We again found clade membership of the virus to be significantly associated with Ct values (N1, N2 primers, *n*=184). Sex was also significant predictor of Ct with males having Ct values 1.09 units lower than females. None of the other predictors were found to be significant in predicting Ct values, which might be driven by a small sample size (Table S1). With ORF1ab primers, clade membership was not significantly associated with Ct values although this is likely due to smaller sample size (*n*=63) (Table S2).

We next investigated which factors impact clinical outcome. To do so, we grouped cases into inpatient (hospitalized) or outpatient (not hospitalized). We then performed a logistic regression with inpatient or outpatient as potential outcomes. As factors predicting the outcome, we considered clade membership, sex, immunocompromised/active cancer, age, week of testing and measured Ct value. The significant predictors for hospitalization were age (*p* = 3.2e-06) and measured Ct value (*p* = 0.012) after Bonferroni Correction for multiple hypothesis testing. Whether a patient was suffering from active cancer and/or was immunocompromised had an estimated odds ratio of 2.9 (0.8-10.8) but was not significant. We did not find any evidence that D614G variant impacts clinical outcome (Fig. 4C). This is consistent with neither variant being enriched significantly in males, immunocompromised/active cancer patients, hospitalized patients, and patients who required intensive care or succumbed due to COVID-19 (Fig. 4D).

## Discussion

The COVID-19 pandemic has greatly impacted lives around the world. As a virus that just recently made the jump into humans, understanding its transmission dynamics and the drivers of its spread are of utmost importance. The emergence of novel, more transmissible strains of SARS-CoV-2 based on an increase in relative frequencies over time has been suggested previously (*23*).

Consistent with trends from other locations around the world (*4*), we find that cases of the spike 614D variant were initially dominating in Washington State, but were later taken over by spike 614G. However, the trends for 614G and 614D cases we observe in Washington State appear to be explained by differences in when action to curb the spread of SARS-CoV-2 were taken on a county level (Figs. 1, 2). The trends in effective reproduction numbers between the two clades 614G and 614D coincide with the different trends in mobility of King County (which includes Seattle) and other areas that experienced substantial spread of SARS-CoV-2. The observed patterns are consistent with initial spread of the 614D clade being largely concentrated in King County, which was then mitigated early on (Fig. 2B). Spread of 614G on the other hand, while present in King County, dominated in other areas of the state and the reduction in the *R*_*e*_ of this variant coincides with a reduction in mobility in these areas, which happened approximately 9 days after King County (Fig. 2B). The spread of SARS-CoV-2 in Yakima County, however, seems to be poorly captured by mobility trends (Fig. S4).

We additionally infer introductions to play a larger role in driving cases of the 614D variant than of the 614G variant. This suggests that differences in the relative frequencies of the two variants are at least in part driven by differences in when and where lineages were introduced into the state. Overall, we find that we can explain the changes in relative frequency of the 614D and 614G variants over time by non-viral factors in absence of intrinsic transmission rate differences. This does, however, not exclude the possibility that such differences exist and have led to the replacement of 614D by 614G in other parts of the world.

We do find evidence for lower Ct values in patients infected with viruses of the 614G variant, which suggests higher viral loads (Fig. 4A,B). This holds, even after including several additional factors, such as the age of a patient and when samples were taken, as potential predictors for Ct values. However, we did not find evidence that D614G has an impact on risk of hospitalization (Fig. 4C,D). The differences in Ct values translates approximately to a 0.42 log_10_ increase in viral load (95% CI: 0.26-0.57 log_10_). This difference might not be large enough to lead to large differences in severity or transmissibility that can be observed in a dataset of this size.

Our findings are broadly consistent with other analyses on the spike D614G substitution. Korber et al. did find evidence of lowered Ct but limited clinical difference for viruses of the 614G clade in Sheffield, UK (*4*). Recent *in vitro* studies show that pseudovirus containing spike protein with 614G substitution exhibits greater infectivity (*5, 8, 9*). Volz et al. suggest increased transmissibility of 614G over 614D in an analysis of thousands of sequences from the United Kingdom (*6*).

Overall, we do find evidence for higher viral loads in individuals with viruses from the 614G clade, which theoretically could impact transmissibility and severity. However, we do not see strong evidence that these differences in Ct values significantly impact the transmissibility or severity of infection with SARS-CoV-2 in the Washington State epidemic.

## Materials and Methods

### Sample collection & testing for SARS-CoV-2

In this manuscript, we analyze 3940 SARS-CoV-2 genomes sequenced from samples collected in Washington State between February and July 2020 as our primary dataset. These samples were pooled across three different channels: UW Virology, WA DOH and SCAN, described below.

For the 1236 UW Virology samples, nasopharyngeal/oropharyngeal swabs were obtained as part of clinical testing for SARS-CoV-2 ordered by local healthcare providers, or collected at drive-up testing sites. RNA was extracted and the presence of SARS-CoV-2 was detected by RT-PCR as previously described using either the emergency use-authorized UW CDC-based laboratory-developed test, Hologic Panther Fusion or Roche cobas SARS-CoV-2 tests (*29*).

For the 2601 WA DOH samples, nasopharyngeal/oropharyngeal/bronchoalveolar/sputum samples were obtained for SARS-CoV-2 clinical testing as requested by submitting healthcare entities. RNA was extracted and the presence of SARS-CoV-2 was detected either via the CDC 2019-nCoV RT-PCR Diagnostic Panel or Applied Biosystems TaqPath COVID-19 Combo Kit.

For the 103 SCAN samples, specimens were shipped to the Brotman Baty Institute for Precision Medicine via commercial couriers or the US Postal Service at ambient temperatures and opened in a class II biological safety cabinet in a biosafety level-2 laboratory. Two or three 650 µL aliquots of UTM were collected from each specimen and stored at 4°C until the time of nucleic acid extraction, performed with the MagnaPure 96 small volume total nucleic acids kit (Roche). SARS-CoV-2 detection was performed using real-time RT-PCR with a probe sets targeting Orf1b and S with FAM fluor (Life Technologies 4332079 assays # APGZJKF and APXGVC4APX) multiplexed with an RNaseP probe set with VIC or HEX fluor (Life Technologies A30064 or IDT custom) each in duplicate on a QuantStudio 6 instrument (Applied Biosystems).

### Viral sequencing & genome assembly

For UW Virology samples, sequencing was attempted on all specimens with Ct < 32 using either a metagenomic approach described previously (*2, 30*), via oligonucleotide probe-capture (*31*), or using an amplicon sequencing based approach (*32*). Libraries were sequenced on Illumina MiSeq or NextSeq instruments using 1×185 or 1×75 runs respectively. Consensus sequences were assembled using a custom bioinformatics pipeline (https://github.com/proychou/hCoV19) that combines de novo assembly and read mapping to generate a per-sample consensus sequence. Consensus sequences were deposited to Genbank and GISAID, and raw reads to SRA under Bioproject PRJNA610428.

For samples from WA DOH and SCAN, sequencing was attempted on all specimens with Ct < 30 using a hybrid-capture approach. RNA was fragmented and converted to cDNA using random hexamers and reverse transcriptase (Superscript IV, Thermo) and a sequencing library was constructed using the Illumina TruSeq RNA Library Prep for Enrichment kit. Using Ct value as a proxy for viral load, samples were balanced and pooled 24-plex for the hybrid capture reaction. Capture pools were incubated overnight with probes targeting the Wuhan-Hu-1 isolate, synthesized by Twist Biosciences. The manufacturer’s protocol was followed for the hybrid capture reaction and target enrichment washes. Final pools were sequenced on the Illumina NextSeq or NovaSeq instrument using 2×150bp reads. The resulting reads were assembled against the SARS-CoV-2 reference genome Wuhan-Hu-1/2019 (Genbank accession MN908947) using the bioinformatics pipeline https://github.com/seattleflu/assembly. Consensus sequences were deposited to Genbank and GISAID.

### Clustering

In order to distinguish between sequences that are connected by local transmission, we cluster all sequences from Washington State together based on their pairwise genetic distance. To do so, we first built a timed tree using sequences from Washington State and from around the world using the Nextstrain pipeline (*3*). Overall, we used 4023 sequences from Washington State and 6028 from the rest of the world. 2601 of all sequences were from the Washington Department of Health, 1236 from the UW Virology Lab, 103 from SCAN. All other sequences were downloaded from the GISAID EpiCoV database (*33, 34*).

We then use a parsimony based approach to reconstruct the locations of internal nodes. To do so, we consider all sequences from Washington State as one location and all sequences from anywhere else on the globe to be from another location. We then reconstruct the internal node locations using the Fitch parsimony algorithm. We consider each group of sequences to be on the same local transmission cluster, if all their common ancestor nodes are inferred to be in Washington State.

### Estimating population dynamics jointly from multiple local outbreak clusters

To estimate the population dynamics of the Washington State outbreak, we use a coalescent approach to infer these dynamics jointly from all known local outbreak clusters. To do so, we model the coalescence and migration of lineages within Washington State as a structured coalescent process with known migration history. The known migration history here is given by the clustering of sequences into local outbreak clusters. The migration events from anywhere outside WA into WA are always assumed to have happened before the common ancestor of all sequences in each local outbreak cluster. How long before this common ancestor time is inferred during the MCMC.

We then infer the effective population size and rates of introductions through time using a skyline type approach. Effective population sizes and rates of introduction are allowed to change at predefined time points. Between these predefined time points where the rates are estimated, the rates are interpolated. This is equivalent to assuming exponential growth or decline between the effective population sizes at these time points.

We then use two different ways to account for correlations between adjacent scaled effective population sizes (*N*_*e*_*τ*). First, we use the classic skyride (*14*) approach where we assume that the logarithm of adjacent *N*_*e*_*τ* is normally distributed with mean 0 and an estimated sigma. Additionally, we use an approach where we assume that the differences ingrowth rates are normally distributed with mean 0 and an estimated sigma. This is equivalent to using an exponential coalescent model with time varying growth rates. We implemented this multi tree coalescent approach as an extension to the Bayesian phylogenetics software BEAST2 (*35*).

The code for the multi tree coalescent is available here (https://github.com/nicfel/NAB) and is validated in Figure S3. We allow the effective population sizes to change every 2 days and the rates of introduction to change every 14 days. The inference of the effective population sizes and rates of introductions is performed using an adaptive multivariate Gaussian operator (*36*), implemented here https://github.com/BEAST2-Dev/BEASTLabs and the analyses are run using adaptive Metropolis coupled MCMC (*37*)

In contrast to backwards in time coalescent approaches, we can consider different local outbreak clusters as independent observations of the same underlying population process using birth death models. We infer the effective reproduction number using the birth-death skyline model (*12*) by assuming the different local outbreak clusters are independent observations of the same process with the same parameters (*13*). We allow the effective reproduction number to change every 2 days. As for the coalescent approach, we assume adjacent effective reproduction numbers to be normally distributed in log space with mean 0 and an estimated sigma. We further assume the becoming un-infectious rate to be 52.3 per year which corresponds to an average duration of infectivity of 7 days (*38*). We allow the probability of an individual to be sampled and sequenced upon recovery to change every 14 days.

### Subsampling of sequence

We analysed the population dynamics in total for 4 different datasets. In the first datasets, we randomly subsample 1500 of the sequences from Washington State, excluding sequences from Yakima County. For the second and third dataset, we distinguish between two different clades we call D and G. The D clade consists of all sequences with an aspartic acid at site 614 of the spike protein. The G clade consists of all sequences with a glycine at this position (visible at https://nextstrain.org/ncov/global?c=gt-S_614). For the 614D datasets, we use the same subsampling procedure as for the above dataset, but with 500 sequences 750 sequences and for the 614G clade. For the dataset from Yakima County, we used 750 randomly subsampled sequences.

### Estimating percentage of introductions of overall new cases

We estimated the relative contribution of introductions compared to local transmission using the coalescent approach introduced here. In addition to the regular assumptions of the coalescent approach that all samples are taken at random from a well mixed population, we assume that differences in effective population size between adjacent time intervals can be used to compute the transmission rate. We then compute the transmission rate as the sum of the growth rate of the effective population size and the becoming uninfectious rate (i.e. we use the relationship *dNe*/*dt* = *transmission rate* − *becoming uninfectious rate* to compute the transmission rate). We assumed an average time of infectiousness of 7 days. Additionally, we assume that *dN*_*e*_/*dt* is independent from the rate of introduction. We then computed the percentage of introductions in overall cases using the rate of introduction and the transmission rate. The rate of introduction can be expressed as the total number of introductions divided by the number of infected in WA, i.e. rate of *introduction* = *nr introductions*/*infected*. The total number of new infections locally can be expressed as *transmission rate* * *infected*, which in turn means that ratio of introductions over local infections can be expressed as *ratio* = (*rate of introduction* * *infected*)/(*transmission rate* * *infected*). From this ratio, we can then compute the percentage of introductions of the overall cases.

We tested that we can retrieve the percentage of introductions from simulations, where we simulated phylogenetic trees using an IR compartmental model with superspreading using MASTER (*39*). We then simulated genetic sequence data using those trees and then inferred the percentage of new cases due to introductions from those sequences (Figs. S9 and S10).

### Chart review

Clinical record review of UW affiliated patients was performed under University of Washington IRB: STUDY00000408. This included patients who visited UW affiliated clinics and patients who were hospitalized at UW Medical Center, both the Montlake and Northwest locations, and Harborview Medical Center. Sex, age, presence of active cancer or immunosuppresive medication, hospital admission, critical care admission, and deceased status was extracted from all charts.

### Factors affecting Ct and clinical outcomes of individuals

R/3.6.2 was used for Ct and clinical record analysis. The code and data cleaned of all patient identifiers is available at: https://github.com/blab/ncov-wa-d614g.

UW Virology used three different primer sets and platforms over the timeframe of the dataset (Fig. S6). Since it is difficult to compare Ct across primer sets, we ran both tests comparing Ct by viral clade and the generalized linear model predicting Ct separately for N1, N2 primers and ORF1ab primers. There were insufficient samples amplified with Egene/RdRp primers for statistical analysis (*n*=20).

We chose to use Wilcoxon Rank Sum Test to compare differences in Ct between viral lineages, and Student’s T-test for comparing differences in age between viral lineages. Age was reported as a decade bin converted into a numerical equivalent, and Wilcoxon Rank Sum Test underestimates differences with duplicate numbers.

For generalized linear models (GLM) of Ct and age, we used a multivariate linear regression of form:

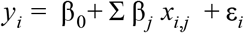

where *y* is the dependent variable (either Ct or age), *β* is the coefficient of the predictor variable, *x* is the predictor variable, and *ϵ* is the residual error. Models were run with the glm package in R (https://www.rdocumentation.org/packages/stats/versions/3.6.2/topics/glm).

UW Virology samples were used to estimate predictors of Ct as SCAN samples were limited in number (*n*=78), and age was not available for WA DOH samples.The predictor variables were amino acid at Spike 614 (binary variable), week since community spread of COVID-19 was reported in Washington (continuous variable), and age of patient (continuous variable).

In the GLM of Ct with only samples from UW Medicine affiliates, we additionally included sex (binary variable), active cancor or immunocompromised (binary variable), hospitalized (binary variable), and required critical care or deceased (binary variable) as predictors of Ct.

To estimate predictors of patient age, we used all SCAN & UW Virology samples with age available (*n*=1172). The predictor variables were amino acid at spike 614 (binary variable) and week since community spread of COVID-19 was reported in Washington (continuous variable).

To estimated predictors of hospitalization if infected with SARS-CoV-2, we used a multivariate logistic regression:

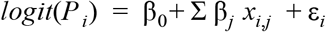

Where *P* is the probability of hospitalization, *β* is the coefficient of the predictor variable, *x* is the predictor variable, and *ϵ* is the residual error. Predictor variables were: week since first sample in dataset (continuous variable), sex (binary variable), active cancer or immunocompromised (binary variable), age in decade (continuous variable), amino acid at Spike 614 (binary variable), and average Ct (continuous variable). To fit the logistic regression, we again used the glm package in R, specifying family as “binomial”. P-values and confidence intervals for risk of hospitalization were adjusted for multiple hypothesis testing using a Bonferroni Correction.

Chi-Squared tests were used to compare proportions of viral lineages by sex, immunocompromised status, clinical outcome (inpatient or outpatient), and severe outcome (critical care or death). P-values were adjusted for multiple hypothesis testing using the Bonferroni Correction.

## Data Availability

Data and code associated with this work are available at https://github.com/blab/ncov-wa-phylodynamics and https://github.com/blab/ncov-wa-d614g. SARS-CoV-2 consensus genome sequences associated with this work have been uploaded to Genbank and the GISAID EpiFlu database and accession numbers are available in supplementary data.

https://github.com/blab/ncov-wa-phylodynamics

https://github.com/blab/ncov-wa-d614g

https://nextstrain.org/groups/blab/ncov/wa-phylodynamics

## Data and materials availability

Sequencing and analysis of samples from the Seattle Flu Study was approved by the institutional review board at the University of Washington (protocol STUDY00006181). Informed consent was obtained for all community participant samples and survey data. Informed consent for residual sample and clinical data collection was waived. Sequencing and analysis of samples from SCAN was approved by the institutional review board at the University of Washington (protocol STUDY00010432). Informed consent was obtained for all community participant samples and survey data. For UW Virology Lab, use of residual clinical specimens was approved by the institutional review board at the University of Washington (protocol STUDY00000408) with a waiver of informed consent. Data and code associated with this work are available at https://github.com/blab/ncov-wa-phylodynamics and https://github.com/blab/ncov-wa-d614g. SARS-CoV-2 consensus genome sequences associated with this work have been uploaded to Genbank and the GISAID EpiFlu database and accession numbers are available in supplementary data.

## Acknowledgements

We gratefully acknowledge the authors, originating and submitting laboratories of the sequences from GISAID’s EpiFlu Database on which this research is based. A full Acknowledgments table is available as supplementary materials. We have tried our best to avoid any direct analysis of genomic data not submitted as part of this paper and use this genomic data as background. We would like to thank Tanja Stadler and Timothy Vaughan for their comments on the phylodynamic analyses.

## Author contributions

NFM designed and performed phylodynamic analyses. CW led individual-level analysis of viral variants. CDF led viral sequencing of SCAN and WA DOH samples. PR led viral sequencing of UW Virology samples. JL, LHM, BP, MR, and ER provided key support for sequence generation and analysis. HX, LS, AA, VMR, NAPL, MH processed and sequenced UW Virology samples. RG, GM, BH, and PD processed WA DOH samples. AA, EB, PDH, KF, MI, KL, TRS, MT, CRW, MB, JAE, MF, BRL, MJR, MT processed SCAN samples. JSD, LMS, HYC, JS, KRJ oversaw sample collection and sequencing. ALG, DAN, and TB oversaw the study. NFM, CW and TB wrote the manuscript. All other authors edited the manuscript.

## Supplemental Materials

**Fig S1.**
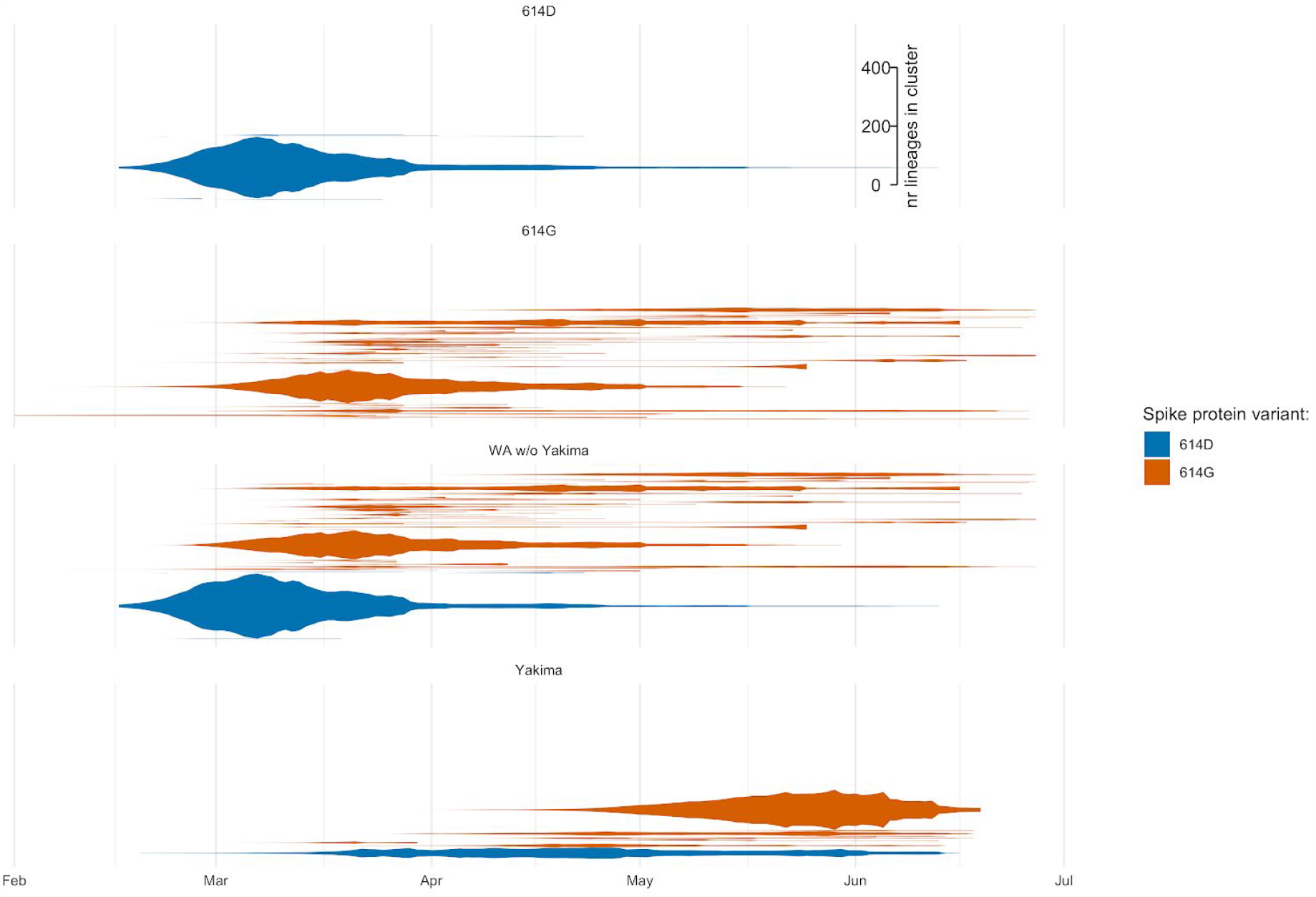
Number of Lineages through time for different local transmission clusters. Here we show the number of lineages in each local transmission cluster (y-axis) over time (x-axis). The different plots show the lineage through time plots for the different datasets analyses here.

**Fig S2.**
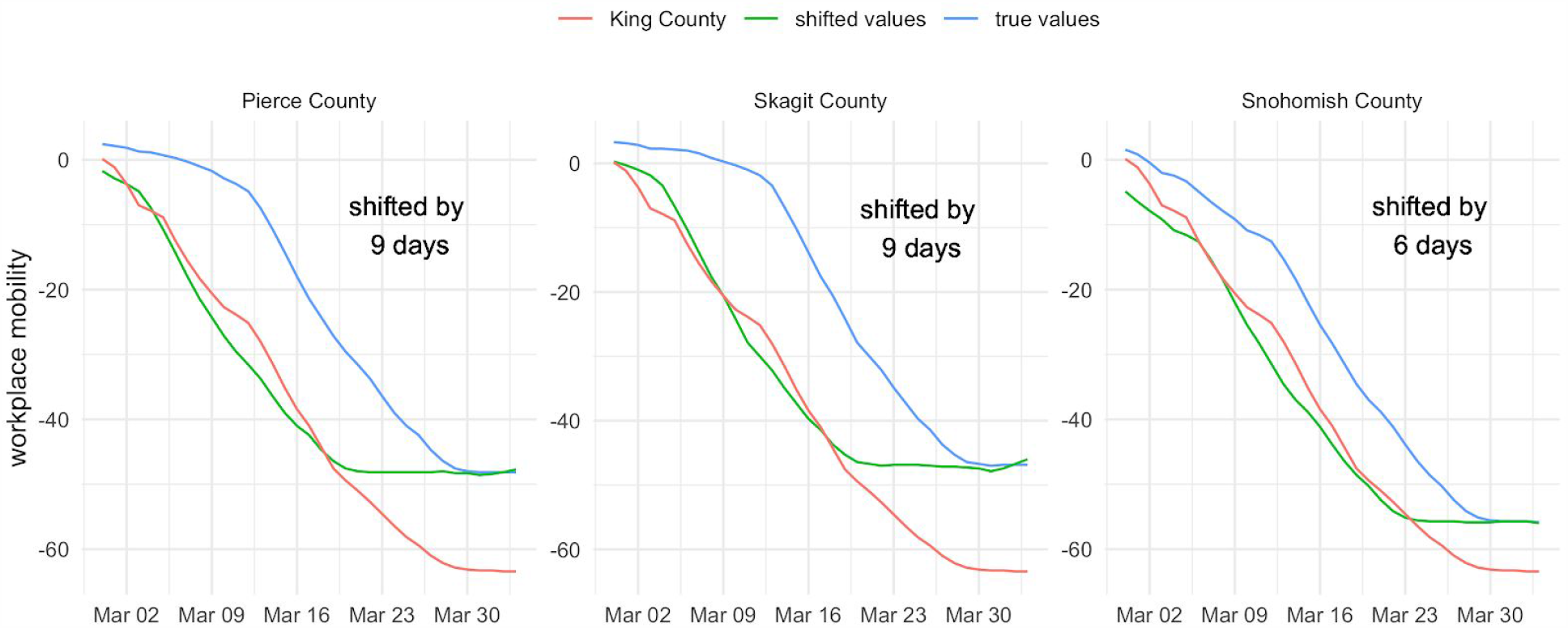
Workplace mobility trends of different counties in Washington State compared to King County. Each plot shows the workplace mobility trend of King County and compares it to either Pierce County, Skagit County or Snohomish County. The red line shows the mobility trend of a county shifted to match the trends in King County. The number of days that the trend line is shifted by is shown in each subplot.

**Fig S3.**
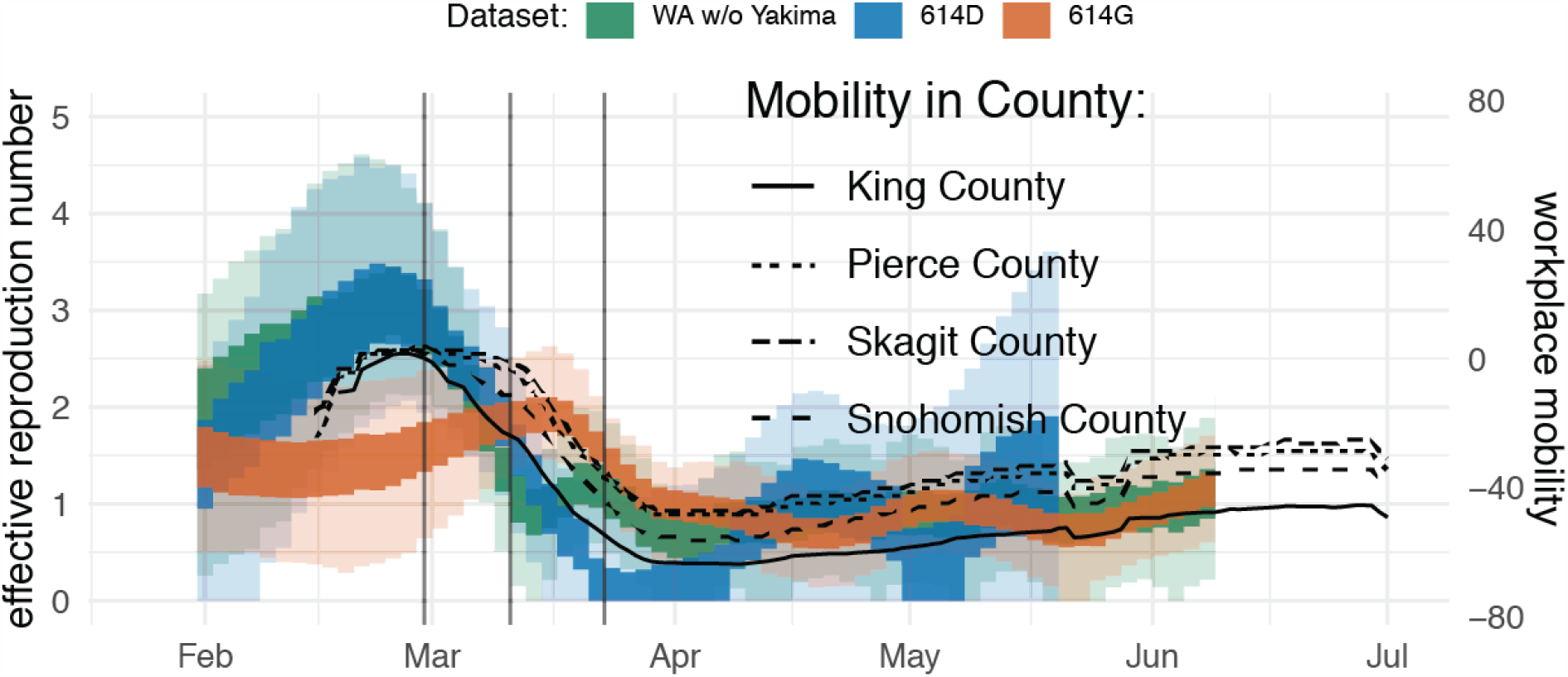
*R*_*e*_ estimates using the coalescent skygrowth model compared to Google mobility data.

**Fig S4.**
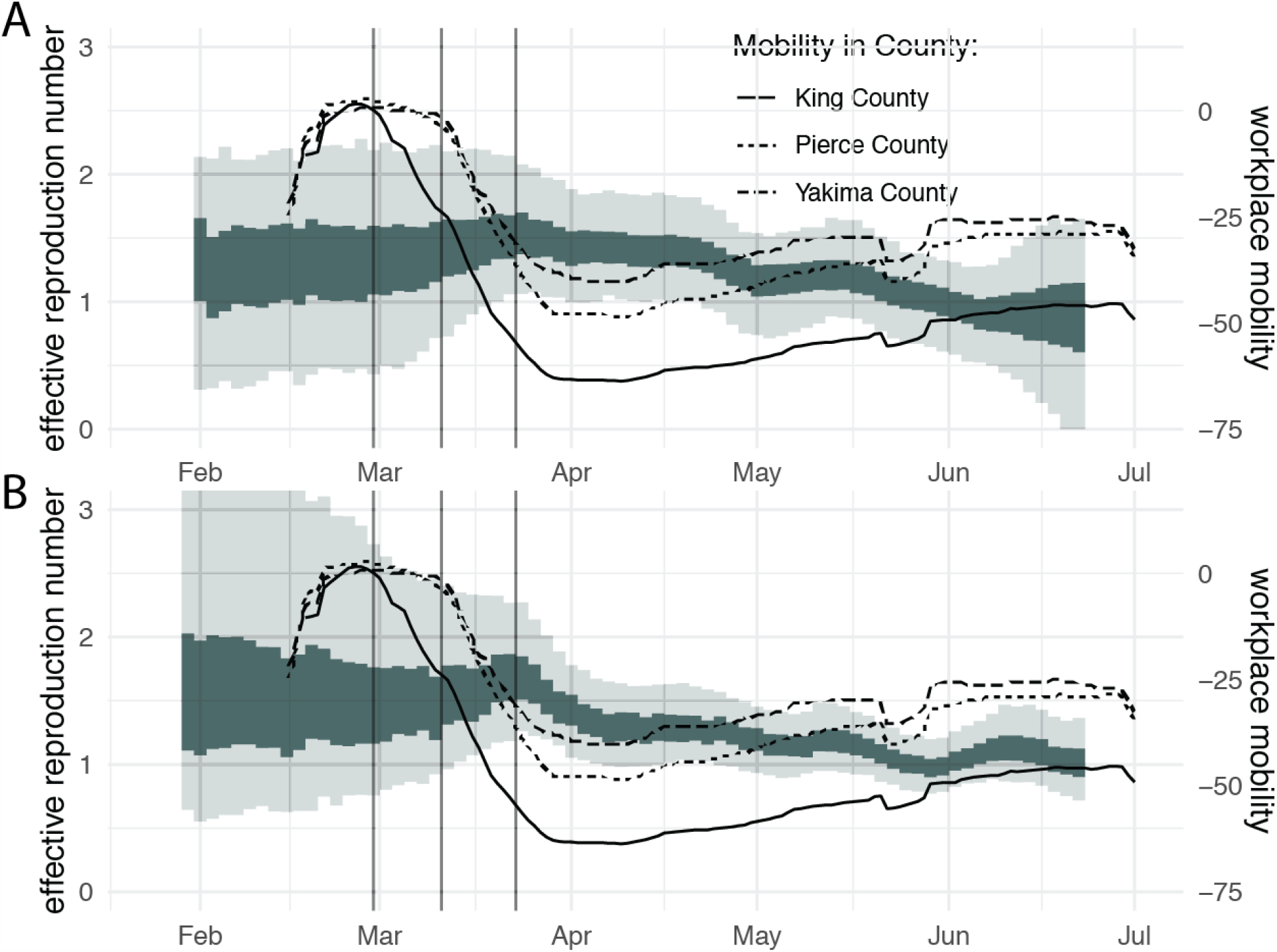
Effective reproduction number and workplace mobility in Yakima County. Here, we show the effective reproduction number estimates over time in Yakima County using the birth-death skyline model (A) and the coalescent skygrowth model (B). The inner band shows the 50% highest posterior density (HPD) interval and the outer band, the 95% HPD interval. Additionally, we compare those estimates to mobility trends in Yakima County and (as a reference) King and Pierce County. The mobility trends are shown as a 7 day moving average.

**Fig S5.**
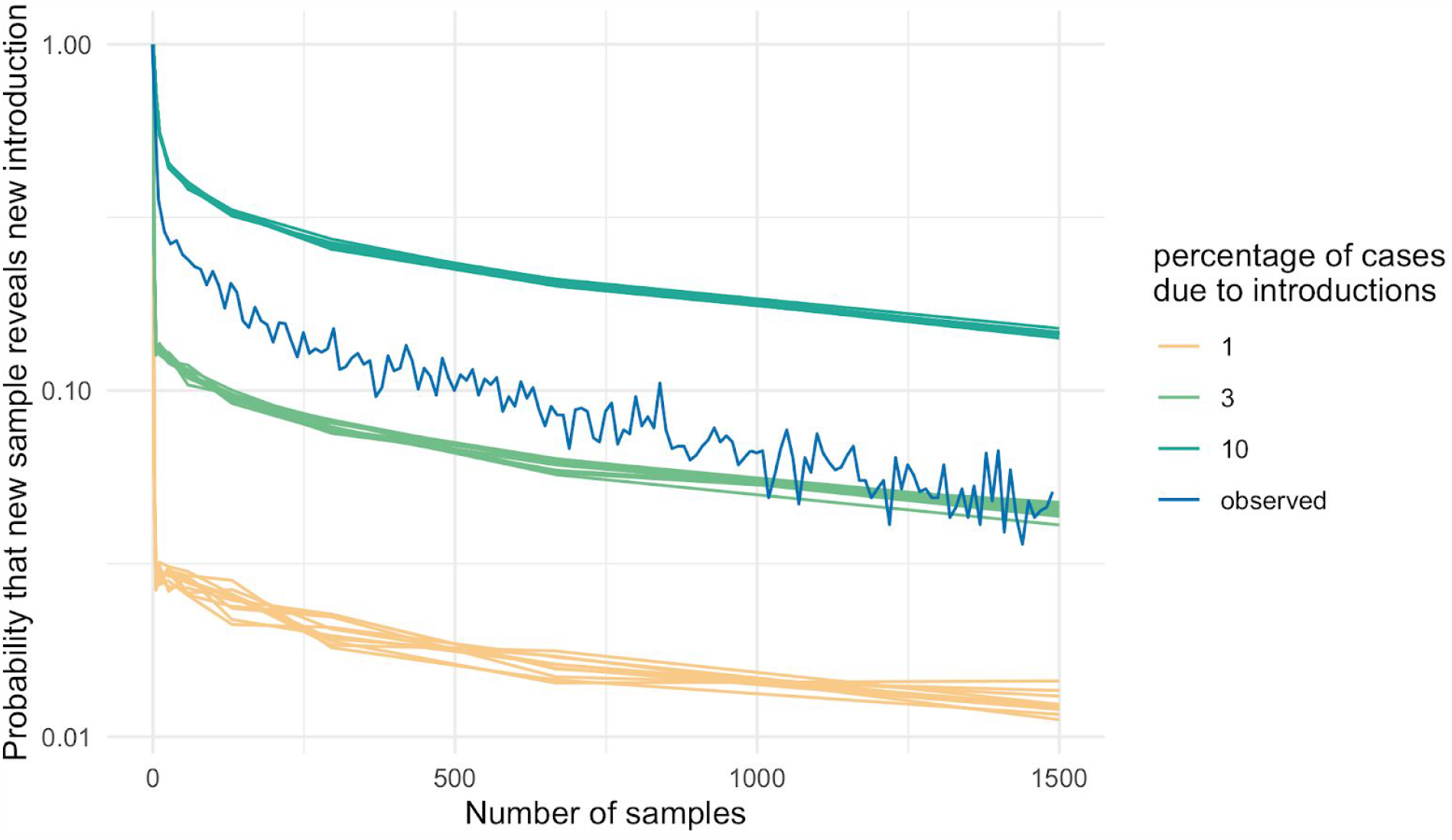
Percentage of introductions due to introductions using cluster size distributions. Here we compare the probability that adding a new sequence to a dataset reveals a new introductions between what we observed empirically and when we simulate clusters using different percentages of introductions. To do so, we randomly chose n samples (x-axis) and then added one additional sample. We then estimate the probability that this additional sample revealed a new introduction (y-axis). We repeated the procedure for simulated clusters with different percentages of introductions in overall cases.

**Fig S6.**
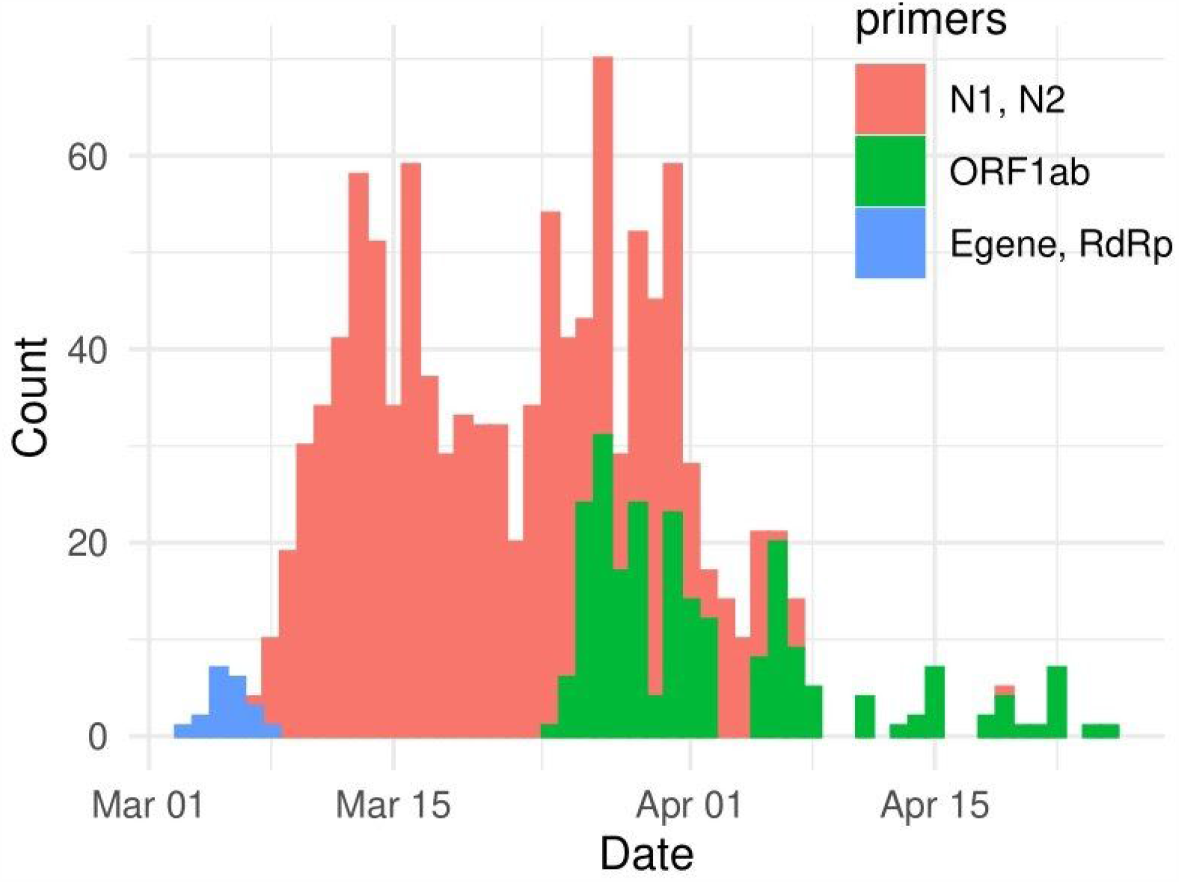
Histogram of primers used by UW Virology across time for samples analyzed for Ct value.

**Fig S7.**
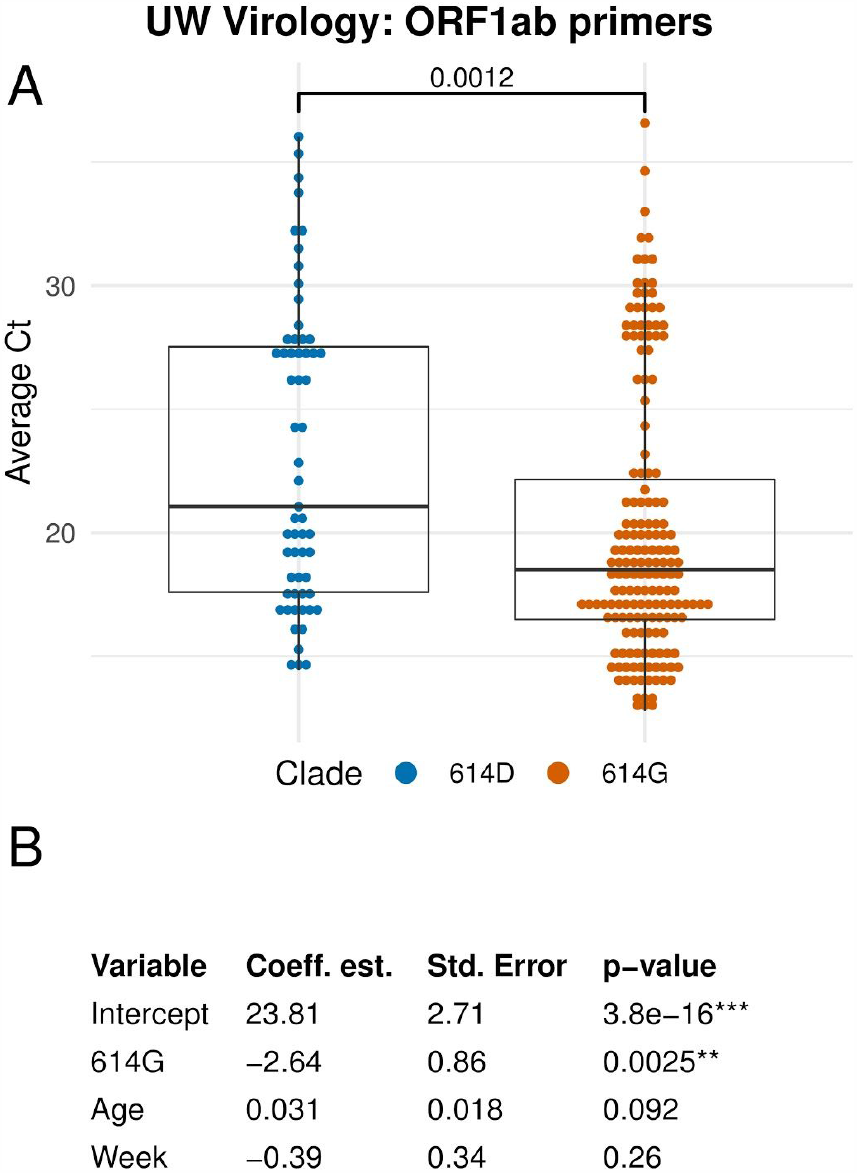
Comparison of cycle threshold (Ct) from ORF1ab primers across SARS-CoV-2 clade. **A** Boxplot of Ct with ORF1ab primers by amino acid at Spike 614. **B** GLM analysis of Ct values from ORF1ab primers using several different predictors.

**Fig S8.**
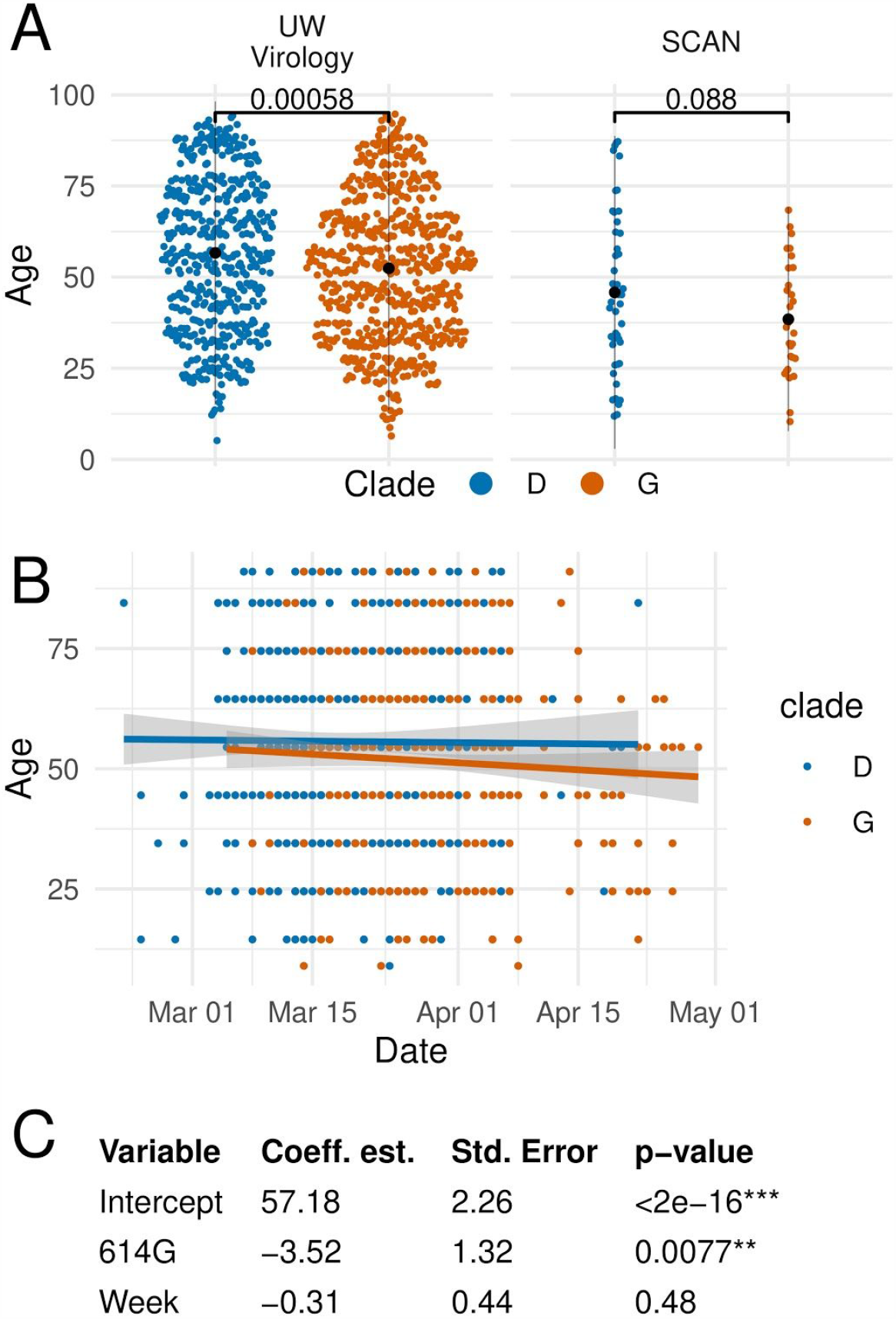
Age of infected individuals by 614D or 614G variant over time. **A** Age of infected individuals in UW Virology and SCAN samples according to D614G variant. Mean age and two standard deviations are shown in black. **B** Age of infected individuals over time partitioned by D614G variant. **C** GLM of patient age predicted by D614G variant and sampling week.

**Fig S9.**
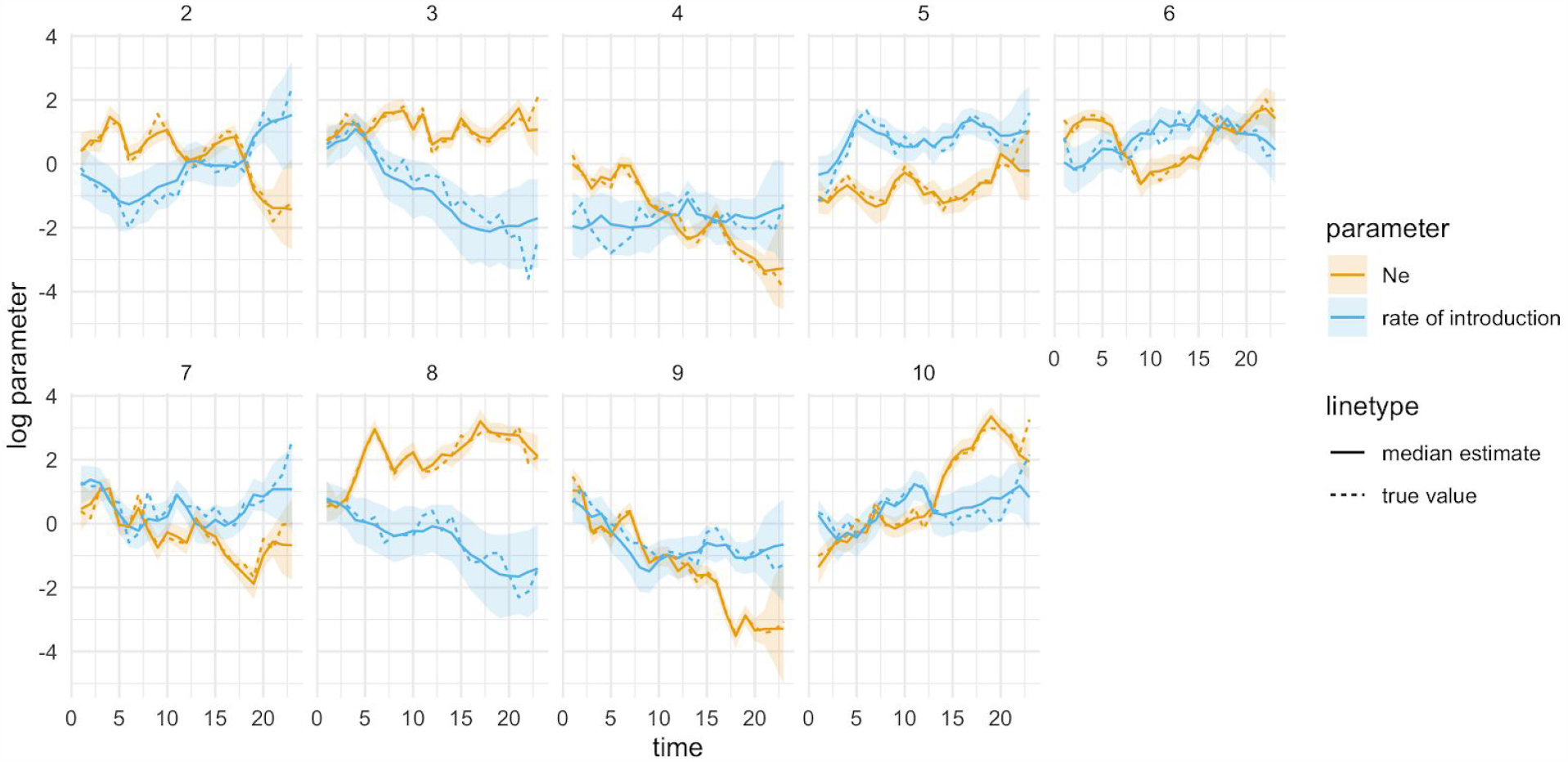
Estimation of effective population sizes and rates of introductions from simulations. Here, we infer effective population sizes and rates of introductions from phylogenetic trees, simulated under the structured coalescent when conditioning on observing a migration history.

**Fig S10.**
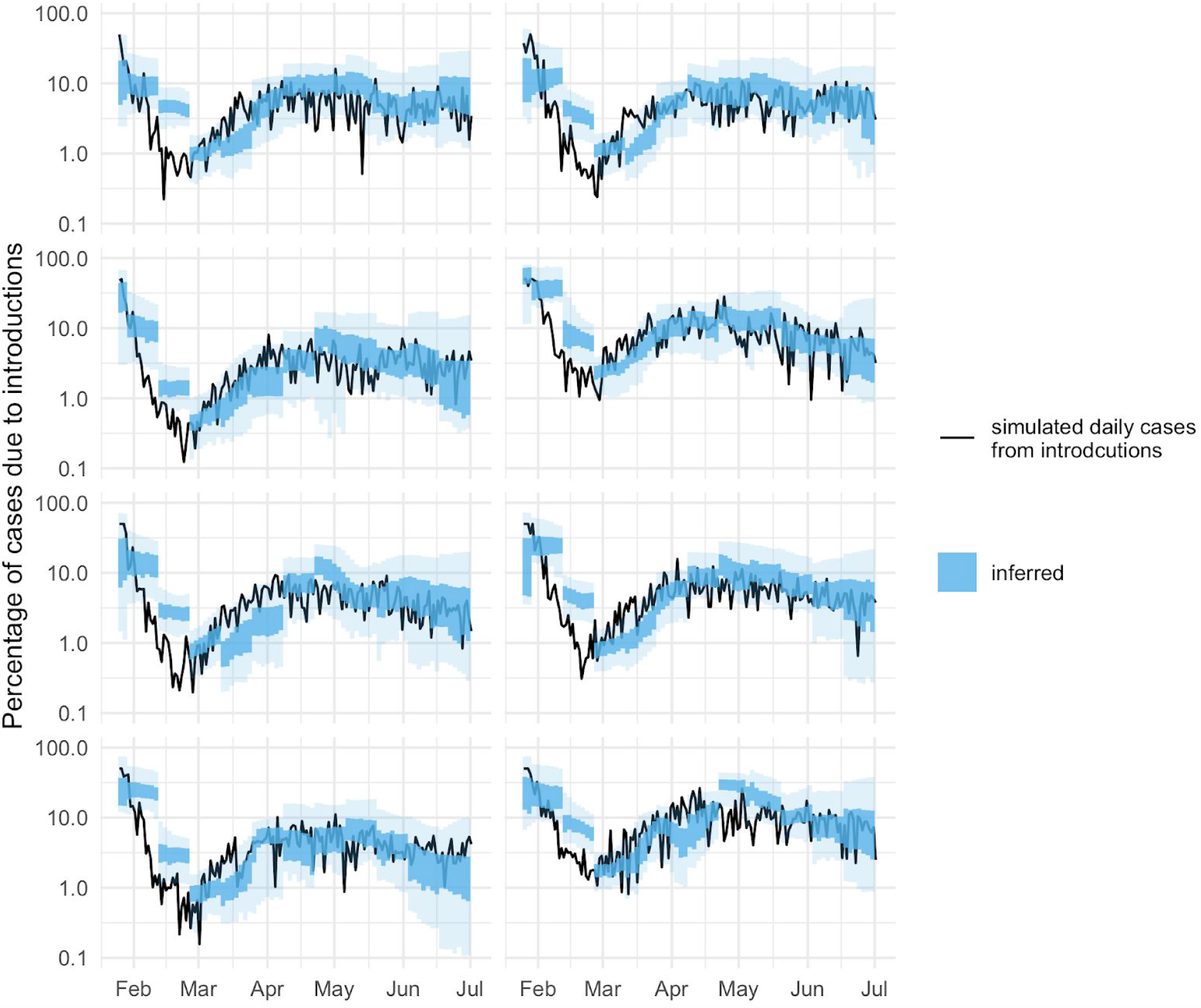
Estimation of effective population sizes and rates of introductions from simulations. Here, we test how well we can retrieve the percentage of new cases due to introductions over time from simulations. To do so, we simulated a local outbreak using a constant rate of introduction. We then simulated genetic sequences and then used the local transmission cluster to estimate the percentage of introductions in blue using the multi tree coalescent.

**Table S1.** GLM of Ct with N1, N2 primers in patients at UW affiliates

| Variable | Coefficient estimate | Std. Error | p-value |
| --- | --- | --- | --- |
| Intercept | 16.00 | 1.45 | <2e-16*** |
| 614G | -1.35 | 0.55 | 0.014* |
| Week since start of WA epidemic | 0.32 | 0.27 | 0.23 |
| Male | 1.09 | 0.48 | 0.023* |
| Age | 0.012 | 0.014 | 0.36 |
| Active cancer or immunocompromised | -0.253 | 0.77 | 0.74 |
| Hospitalized | 1.22 | 0.77 | 0.11 |
| Critical care and/or deceased | -0.65 | 0.98 | 0.51 |

**Table S2.** GLM of Ct with ORF1ab primers in patients at UW affiliates

| **Variable** | **Coefficient estimate** | **Std. Error** | **p-value** |
| --- | --- | --- | --- |
| Intercept | 23.13 | 6.39 | 6.6e-04*** |
| 614G | 1.07 | 1.92 | 0.58 |
| Week since start of WA epidemic | -0.75 | 0.72 | 0.30 |
| Male | 0.76 | 1.76 | 0.67 |
| Age | 0.013 | 0.044 | 0.76 |
| Active cancer or immunocompromised | 2.18 | 2.92 | 0.46 |
| Hospitalized | 1.22 | 0.77 | 0.11 |
| Critical care and/or deceased | -2.96 | 2.99 | 0.33 |

